# Spontaneous Clearance of Vertically Acquired Hepatitis C Infection: Implications for Testing and Treatment

**DOI:** 10.1101/2021.09.29.21264077

**Authors:** A E Ades, Fabiana Gordon, Karen Scott, Jeannie Collins, Claire Thorne, Lucy Pembrey, Elizabeth Chappell, Eugènia Mariné-Barjoan, Karina Butler, Giuseppe Indolfi, Diana M Gibb, Ali Judd

**Affiliations:** Population Health Sciences, University of Bristol Medical School, Whatley Road Bristol BS8 2PS; MRC Clinical Trials Unit, University College London, 90 High Holborn, London WC1V 6LJ; Population, Policy and Practice Research and Teaching Department, UCL Great Ormond Street Institute of Child Health, 30 Guilford Street, London WC1N 1EH; London School of Hygiene and Tropical Medicine, Keppel Street, London WC1E 7HT; Université Côte d’Azur, Public Health Department. Centre Hospitalier Universitaire de Nice, Rte St Antoine de Ginestière BP 3079. 06202 Nice cedex 2; Children’s Health Ireland at Crumlin and Temple Street, Dublin, Ireland; Meyer Children’s Hospital and Department Neurofarba, University of Florence, Viale Gaetano Pieraccini, 24, 50139 Firenze FI, Italy

**Keywords:** Hepatitis C virus, HCV, vertical transmission, spontaneous clearance, over-treatment

## Abstract

**Background:** Current guidelines recommend that infants born to women with hepatitis C (HCV) viremia are screened for HCV antibody at age 18 months, and if positive, referred for RNA testing at 3 years to confirm chronic infection. This policy is based in part on analyses suggesting 25%-40% of vertically acquired HCV infections clear spontaneously within 4-5 years.

**Methods:** Data on 179 infants with RNA and/or anti-HCV evidence of vertically acquired viraemia (single PCR+) or confirmed infection (2 PCR+ or anti-HCV beyond 18 months) in three prospective European cohorts were investigated. Ages at clearance of viremia and confirmed infection were estimated taking account of interval censoring and delayed entry. We also investigated clearance in infants in whom RNA was not detectable until after 6 weeks.

**Results:** Clearance rates decline rapidly over the first 6 months. An estimated 90.6% (95%CrI: 83.5-95.9) of viremia cleared by 5 years, most within 3 months, and 65.9% (50.1-81.6) of confirmed infection cleared by 5 years, at a median 12.4 (7.1-18.9) months. If treatment began at age 6 months, 18 months or 3 years, at least 59.0% (42.0-76.9), 39.7 (17.9-65.9), and 20.9 (4.6-44.8) of those treated would clear without treatment. In seven (6.6%) confirmed infections, RNA was not detectable until after 6 weeks, and in 2 (1.9%) not until after 6 months. However, all such cases subsequently cleared.

**Conclusions:** Most viraemia clears within 3 months, and most confirmed infection by 3 years. Delaying treatment avoids but does not eliminate over-treatment and should be balanced against loss to follow-up.

**Key points:** Based on a re-analysis of the largest purely prospective dataset assembled so far, 66% (50-82) of confirmed vertically acquired HCV clears spontaneously by age 5 years, rather than the 25-40% assumed in guidelines.

---

The development of highly effective direct acting antiviral (DAA) treatment for HCV[1, 2] has raised the possibility that vertically acquired infection, which occurs in 5%-6% of deliveries to HCV-RNA positive mono-infected women,[3, 4] could be prevented or treated.[5] Screening of women in pregnancy, followed by treatment in late pregnancy or postnatally, as well as treatment of infected infants, have both been considered,[2, 6] and could contribute substantially to the WHO targets to eliminate HCV by the year 2030.[7] However treatment in pregnancy is not currently recommended in spite of good safety profiles.[8] DAA treatment in children is approved from age 3 years in the European Union and the USA, and WHO recommendations for lower and middle income countries (LMICs) are under review.[9-12] Work on pediatric formulations and pharmacokinetic studies is ongoing.

Diagnosis of HCV infection in infants exposed in utero is based largely on the presence of HCV antibodies (anti-HCV) at 18 months.[10-13] Those who are anti-HCV positive are referred for RNA testing at age 3 years to confirm chronic infection.[10]

The policy of deferred testing and treatment is supported by the assumption in recent reviews and guidelines that 25%-40% of vertical infections clear spontaneously by age 5 years,[10-12, 14] as indicated by repeatedly negative HCV-RNA and/or clearance of anti-HCV. However, these estimates are based on studies in which children have not been tested regularly from birth and which may miss early clearance. In other studies infants were recruited retrospectively and inclusion depended on a period of sustained follow-up, potentially resulting in selective inclusion of non-clearers. The 25%-40% clearance estimate derives from the European Pediatric HCV Network (EPHN) study,[15] which included 40% retrospectively recruited children. See Supplementary Materials for a review of the literature.

Our objective was to obtain unbiased estimates of spontaneous clearance rates by using a large, purely prospective dataset of infants born to HCV infected women, followed from birth, using statistical methods appropriate for the interval censoring and left truncation in the data. Interval censoring refers to the fact that the age at clearance is never observed, only the ages at the last positive HCV-RNA test and the first negative one. Left truncation, also known as “delayed entry”, occurs if the first HCV RNA positive is not at or shortly after delivery: this would result in early clearance being missed. Failure to account for either of these features will tend to underestimate clearance rates.

Our primary focus is clearance of infections that have been confirmed by at least one additional positive RNA test, at least 6 weeks and usually over 3 months apart. However, we also investigate clearance of viremia, which includes cases where evidence for vertically-acquired HCV is based on a single positive RNA result. Previous authors [16] have referred to these as transient infections.

Based on our estimates of clearance, we make projections about the possible extent of “over-treatment” of confirmed infections if treatment was initiated at 6, 18, or 36 months. In addition, we estimate the frequency of late appearance of detectable RNA in infected infants, and clearance in this group.

## METHODS

### Sources of data

We had direct access to two datasets held at the Great Ormond Street Institute of Child Health, University College London. These were the European Pediatric HCV Network (EPHN),[3, 17, 18] and a study by the British Pediatric Surveillance Unit (BPSU) including data from three maternity hospitals in Dublin.[19] We approached the investigators of 17 other published prospective studies which followed over 100 HCV-infected pregnant women and their infants. One of these, the ALHICE study (Alpes-Maritimes, Languedoc, Haute Garonne Infection C chez l’Enfant)[20] agreed to provide patient-level data. The risk factor distributions, periods of recruitment, and follow-up schedules are summarized in Supplementary Materials.

### Definitions of infection and clearance

*Confirmed infection:* detection of RNA on at least two occasions, or of anti-HCV after 18 months. *Viremia:* as for confirmed infection, but only one positive RNA required. C*learance:* To qualify as a clearer the child’s last RNA test had to be negative, or in the absence of an RNA test, the last HCV antibody test must be negative. In view of the high specificity of these tests[21, 22] only a single marker of clearance was required, either a negative RNA or a negative anti-HCV test. Clearance was considered to have occurred before the date of the first of two consecutive negative markers, and after the immediately preceding positive RNA test. If there was only one marker of clearance, it was considered to have occurred before that marker. A two-marker criterion of clearance was included as a sensitivity analysis. See Supplementary material for further details and supporting information.

### Statistical methods

Restricted cubic spline time-to-event (“survival”) models[23] were fitted to both the viremia and confirmed infection datasets, with clearance as the end-point, taking interval censoring and delayed entry into account. Cubic splines are flexible models that can fit the data smoothly without assuming any particular form. Estimation was carried out by Bayesian Markov Chain Monte Carlo. Parametric models with cure parameters were used as sensitivity analyses. Further statistical details are given in Supplementary materials.

Based on the estimated clearance curve for confirmed infection, we calculated the proportion of all infected children would be potentially “over-treated” if treatment was begun at 6, 18, or 36 months (Public Health perspective), in the sense that they would have cleared spontaneously in the absence of treatment. We also calculated the proportion of *treated* infection that would have cleared anyway if treatment began at those ages (Clinical perspective). (See Supplementary materials)

Finally, we identified infants with at least one positive RNA test, but whose first RNA test was negative. Those who met the definition of confirmed infection, and who were still RNA negative until after 6 weeks of age, were selected for a record review.

### Sensitivity analyses

Sensitivity analyses were carried out to assess robustness of results and conclusions to the form of the survival model. A further sensitivity analyses explored the possibility that cases of viremia based on single RNA positives might be false positives (See Supplementary materials). A third sensitivity analysis looked at a dataset with a more restricted definition of clearance requiring two markers.

## RESULTS

### Time to clearance of infection

106 infants met the definition of Confirmed Infection (Table 1), and an additional 73 of viremia. In all, 87 of the total 179 infants showed evidence of clearance. A very small minority of infants tested RNA positive in the first 3 days of life, and in over 50% clearance could not be observed until after 3 months, indicating a considerable potential for clearance before infection was detected, and still more before infection could be confirmed by a second positive RNA test.

**Table 1.** Numbers meeting the definitions for confirmed infection and viraemia by age at first HCV-RNA positive test.

| Age at first RNA +ve | Confirmed infection |  |  | Viraemia |  |  |
| --- | --- | --- | --- | --- | --- | --- |
|  | Non-clearers | Clearers | Total (%) | Non-clearers | Clearers | Total (%) |
| **0 - 3d** | 6 | 3 | 9 (8.5) | 10 | 13 | 23 (12.8) |
| **3d - 6w** | 14 | 8 | 22 (20.8) | 15 | 24 | 39 (21.8) |
| **6w - 3m** | 20 | 5 | 25 (23.6) | 25 | 14 | 39 (21.8) |
| **3m - 6m** | 18 | 9 | 27 (25.5) | 25 | 20 | 45 (25.1) |
| **Over 6m** | 12 | 11 | 23 (21.6) | 17 | 16 | 33 (18.4) |
| **TOTAL** | 70 | 36 | 106 (100) | 92 | 87 | 179 (100) |

Viremia cleared very rapidly, mostly within three months (Figure 1, panel A), with 79.6% (95%CrI: 69.0-89.7) clearing by 12 months, and 90.6% (83.5-95.9) by 5 years. With confirmed infection 57.3% (44.7-68.8) cleared by 3 years and 65.9% (50.1-81.6) five years. Among those who clear in 5 years the median age at clearance is 12.4 months (7.1-18.9). The rate at which clearance occurs, the risk per month, declines markedly over time (Figure 1 Panel B, note the log scale) although the rate of decline is less with confirmed infection.

**Figure 1.**
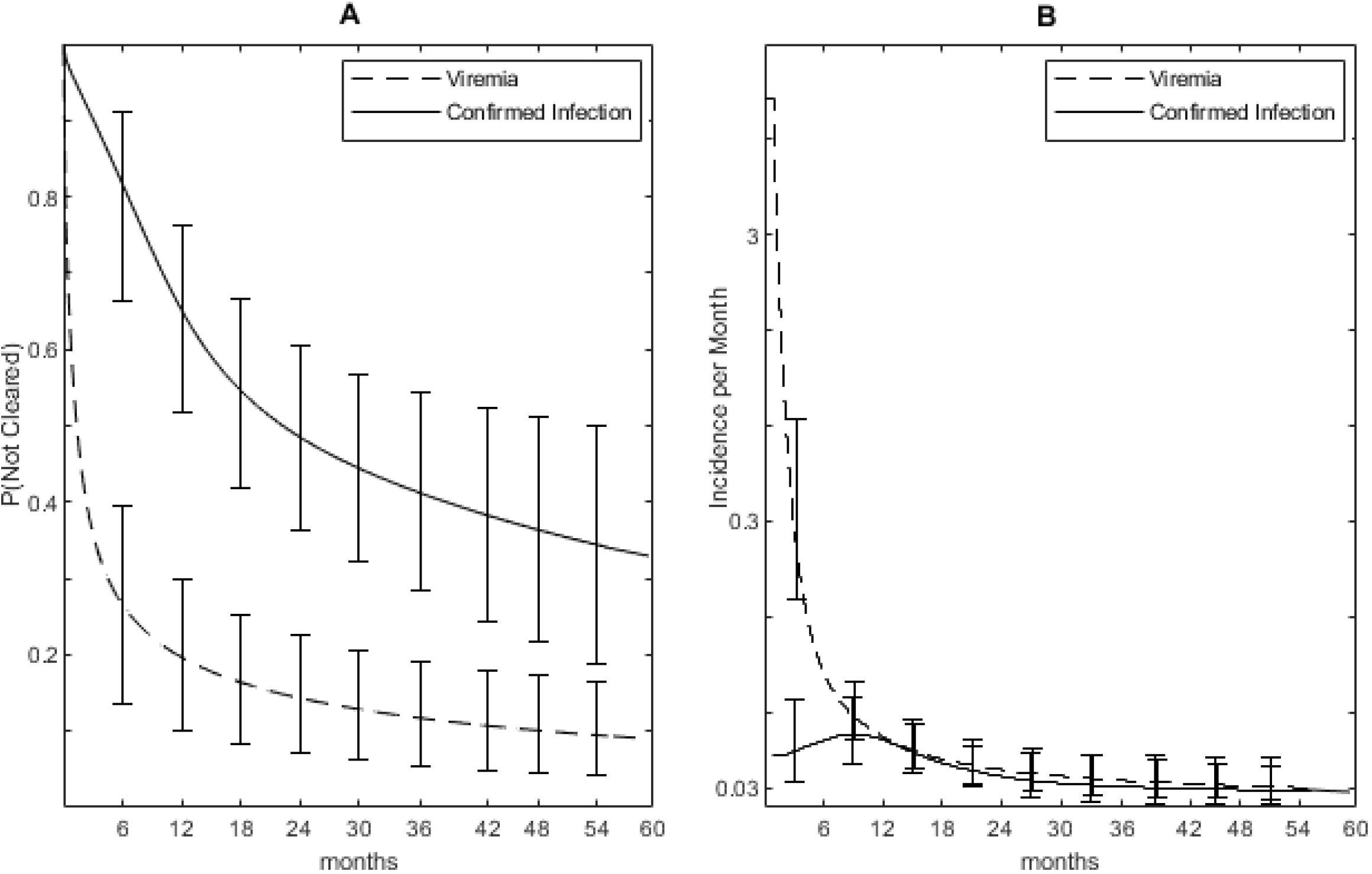
**Panel A**: Proportion remaining infected by age (months) with 95% credible error bars. **Panel B**: Rate of clearance per month with 95% credible error bars on a log scale. Solid line confirmed infection, dotted line viremia.

### Risk of “over-treatment”

Table 2 considers scenarios in which treatment is administered at different ages. Based on estimates of clearance of confirmed infection, we estimated the proportion who would be potentially “over-treated” – i.e. who would clear spontaneously in the absence of treatment. Results are presented for two scenarios: either assuming that no further spontaneous clearance occurs after age 5 years, or that 25% of those still infected at 5 years will eventually clear. With a public health perspective, assuming no further clearance after age 5, then 74.3 (54.3-87.6), 33.6 (17.0-51.8) and 12.8 (3.8-23.1) of all confirmed infections would be overtreated if treatment was begun at 6, 18, and 36 months respectively. From a clinical perspective and under the same assumptions, at 6, 18, and 36 months 59.0 (42.0-76.9), 39.7 (17.9-65.9), and 20.9 (4.6-44.8) of *remaining* infections – i.e. of the infections that would be actually treated - would be expected to clear without treatment.

**Table 2.** Projected proportion “over-treated” in that they would be expected to clear without treatment, assuming: no further clearance of infection remaining at age 5 years, or 25% further clearance. (A) Public Health perspective: estimated proportion of *all confirmed infections* that would be potentially overtreated, if treatment began at each age. (B) Clinical perspective: estimated proportion of *remaining confirmed infections* over-treated at each age. Estimates based on clearance of confirmed infection (Figure 1 Panel B).

| Age Starting Treatment, months | (A). Public health perspective<br>% of all infections that will clear |  | (B). Clinical perspective<br>% of remaining infections that will clear |  |
| --- | --- | --- | --- | --- |
|  | No clearance after 5 years | 25% more clearance after 5 years | No clearance after 5 years | 25% more clearance after 5 years |
|  | % (95%CrI) | % (95%CrI) | % (95%CrI) | % (95%CrI) |
| **6** | 74.3 (54.3-87.6) | 77.2 (58.9-89.1) | 59.0 (42.0-76.9) | 69.2 (56.5-82.7) |
| **18** | 33.6 (17.0-51.8) | 41.6 (27.7-56.5) | 39.7 (17.9-65.9) | 54.8 (38.4-74.4) |
| **36** | 12.8 (3.8-23.1) | 23.1 (15.2-32.0) | 20.9 (4.6-44.8) | 40.7 (28.5-58.6) |

If it is assumed that 25% of those remaining infected at 5 years will eventually clear, the extent of potential over-treatment is proportionately higher (Table 2).

### Time to detectable RNA

A record review of the 7 babies (6.6% of the 106) in whom confirmed infection was not detected until after 6 weeks is shown in Table 3. In most cases there were more than 2 positives, and more than 2 final negative tests, so it is unlikely that either the diagnosis of infection or of clearance are due to diagnostic errors. However, in 5/7 cases there was only a single initial negative test over 6 weeks, and there is a possibility these were false negatives.

**Table 3.** Record review of 7 infants meeting the definition for Confirmed Infection (at least two positive RNA tests) who were initially RNA negative, and who remained RNA -ve until after 6 weeks. Age ranges in months

| Case | Age last RNA -ve | No. of initial RNA -ves > 6w | Age first RNA +ve | Number of RNA +ves | Age last RNA+ve | Number of final RNA -ves | Age last antibody +ve | Age first antibody -ve | Ever breastfed | Mode of delivery | Mother HIV | Mother PCR | Infant HIV | Highest ALT, IU / L | Hepatomegaly |
| --- | --- | --- | --- | --- | --- | --- | --- | --- | --- | --- | --- | --- | --- | --- | --- |
| 1 | 0-3 | 1 | 6-9 | 2 | 9-12 | 3 | 9-12 | 12-15 | Yes | EC-S | Neg | Pos | No | 40-80 | No |
| 2 | 0-3 | 1 | 12-15 | 2 | 15-18 | 2 | 0-3 | 18-24 | Yes | EC-S | Neg | Neg | No | 40-80 | No |
| 3 | 0-3 | 2 | 3-6 | 3 | 15-18 | 3 | 3-6 | 9-12 | Yes | Other | Neg | Pos | No | 40-80 | No |
| 4 | 6-9 | 1 | 6-9 | 4 | 36-48 | 1 | 36-48 | - | No | EC-S | Neg | NK | No | >80 | Yes |
| 5 | 6-9 | 2 | 9-12 | 3 | 24-36 | 3 | 9-12 | 12-15 | No | Other | Neg | Pos | No | <40 | No |
| 6 | 0-3 | 1 | 3-6 | 5 | 12-15 | 2 | 0-3 | 3-6 | No | EC-S | Neg | NK | No | <40 | No |
| 7 | 0-3 | 1 | 0-3 | 3 | 24-36 | 4 | 15-18 | - | No | Other | Neg | Pos | No | >80 | No |

EC-S: Elective Caesarean Section. NK: not known

In 4 cases, RNA was not detected until after negative findings at age 3 months, and in 2 cases after 6 months. However, all 7 cases eventually became RNA negative between 12 and 44 months, 6 meeting the criteria for confirmed clearance. Two, however, were anti-HCV positive at 15-18 and 36-48 months respectively and had experienced ALT levels over 80 IU/l. The risk factors for the 7 infants were unremarkable; 3/7 (43%) had breastfed, compared to 28% in the whole EPHN cohort (Supplementary Materials Table B1).

### Sensitivity analyses

For both viremia and for confirmed infection, there were no material differences between the preferred cubic spline model and the three parametric survival models in either model fit, median time to clearance, or estimated age at clearance (Supplementary materials Tables E1, E2). The impact of specificity-adjustment on viremia clearance was also negligible (Supplementary Table E2). Somewhat less clearance was observed with the stricter definition of clearance requiring 2 markers, but little impact on estimated risks of “over-treatment” if treatment was given at age 36 months (Supplementary materials Table E2).

## DISCUSSION

Based on data from three cohort studies following prospectively recruited vertically infected infants from birth, we estimate that 90.6% (95%CrI 83.5-95.9) of vertically acquired viraemia clears by 5 years, mostly before 3 months. 83.0% (73.8-91.5) has already cleared by 18 months.

These results are entirely consistent with studies in which children are tested at birth and in which a single RNA test is taken as indicative of potential infection (Supplementary Materials Table A1). The largest study of this sort[16] reported 75% clearance by 1.8 years. In addition, all those infected who were RNA positive at delivery – nearly 30% of the total infected – had cleared by 4 months. Because our analysis takes delayed entry into account, it recovers estimates that are similar to these studies, even though the testing at delivery was not always undertaken in the three cohorts studied here.

The results obtained on confirmed infection are those most relevant to inform management policy, as confirmatory testing would be routinely required before treatment was considered. In this scenario, 56.0% (44.7-69.8) clear by age 3 years, and 65.9% (50.1-79.6) by 5 years. These estimates are substantially higher than the 25-40% clearance rates that are commonly cited,[5, 12, 24] which are partly based on datasets in which infants have either not been tested and followed from birth, and which have therefore missed cases of early clearance, or studies which have included retrospectively recruited infants. Studies in which clinics are asked to both retrospectively include children and then follow them prospectively may be especially vulnerable to bias, unless they include all those who have already been discharged or otherwise lost to follow-up at the time the clinic joins the study.

The marked fall in clearance rates with age, which has not been reported previously, further underscores the danger of ignoring delayed entry in studies where infected infants are first tested *after* the period of high clearance rates is over.

Sensitivity analyses indicate that our findings are robust to alternative types of time-to-event model, and to potential false positive RNA results. Lower rates of clearance were seen when clearance was based on a two-marker definition. However, although a two-marker definition of clearance is more relevant in a clinical setting, it is likely that these analyses are biased by mis-specifying single negative RNA tests as false negatives and hence as censored observations. We estimated the Negative Predictive Value of RNA tests to be over 98%, consistent with previous research;[21, 22] false negative RNA results would therefore be highly infrequent.

Current guidance recommends testing for HCV antibodies at 18 months and referral of positives for RNA testing at 3 years to confirm chronic infection prior to treatment.[10] A focus on treatment of chronic infection is reasonable at this time when no treatments are available under age 3. However, this strategy may not be sustainable longer term. Loss to follow-up between 50% and 80% has been reported in a number of US studies,[25-27] with less than 50% of children followed to 18 months.[28] Higher follow-up rates should be achievable in well-resourced countries with centralized health systems, but follow-up to even 6 months may be seen as ambitious in lower and middle income countries, where delayed treatment may in many cases result in no treatment, with an attendant risk of liver disease and onward transmission.[5]

HCV-RNA testing between 2 and 6 months, which correlates well with results at 18 months,[10] can also be useful as negative findings, which would be expected in 95% of those exposed, usually rule out chronic infection.[10] A balance must be found between early treatment with its risk of over-treatment and deferred treatment with its risk of no treatment, and this balance may depend on the health system as well as the underlying epidemiology and burden of maternal infection.

While delaying treatment reduces the extent of over-treatment, delay to even 36 months may not eliminate it. Even if we assume no further clearance after age 5 years (Table 2), our projections were that 74.3%, 33,6% and 12.8% of *all* confirmed infections, and 59.0%, 39.7% and 20.9% of *all treated* infections, would be clear spontaneously if treatment was to begin at age 6, 18, and 36 months respectively. The figures are higher if more clearance occurs after age 5 years. Clinic-based studies (Supplementary materials Table A3) suggest that 2.3%-29% after 5 years but these estimates cannot be relied on for reasons given above. More data on rates of clearance after age 3 years would be valuable.

One advantage of early treatment is that it may require much smaller doses, and it has even been suggested that a short course of DAA immediately after birth, and before infection could be confirmed, could prevent vertical transmission.[29] Otherwise, six months is probably the earliest age at which treatment of confirmed infections could be considered as RNA tests at 2 to 3 months could be fitted in with vaccination schedules while allowing for confirmatory tests at 6 months. It has been claimed that infants with negative RNA results at two months do not require further testing,[26] but this is counter-indicated by previous analyses[22] and by isolated reports of late-appearing RNA throughout the literature.[30, 31] In our study 7 of 106 confirmed infected babies were still RNA-ve after 6 weeks, including 2 after 6 months, although all eventually cleared. Perhaps of more concern are reports of the *re-*appearance of infection after clearance of RNA,[32] or *re-*appearance of antibody after clearance of maternal antibody.[16] These may be postnatally-acquired infections: intra-familial transmission can occur but is unusual[33] and breastfeeding is not believed to be a risk factor.[3, 9] In any case, if earlier diagnosis is to be considered, sufficient time must be allowed for vertical infections that do not clear to become manifest, and more information is required on when this might be.

Given the remaining uncertainties about rate of clearance and possible over-treatment, and the difficulties surrounding diagnosis and follow-up, it could be argued that attention should focus on treatment to prevent transmission in pregnancy, rather than treatment of pediatric infection.

Although this is the largest purely prospective dataset assembled to date, there are important limitations. Foremost is the historical nature of our dataset, collected between 1994 and 2004, when HCV-RNA testing was less accurate and less standardized. This emphasizes the need for additional data. A further difficulty is that confirmed infection is not a well-defined construct: estimates of both the VT rate and the extent of clearance both depend on the precise details of the definition and on the intensity of the follow-up schedule, as noted previously:[16, 30] more frequent testing will result in higher VT rates and more clearance. There have been very few studies investigating both vertical transmission of HCV and its subsequent clearance in the exact same cohort, and therefore no estimates of what vertical transmission rates might be “net” of clearance at different ages. This is the subject of a companion paper based on the same three cohorts.

This paper has clarified the extent of spontaneous clearance and has set out for the first time the statistical and study design considerations required to obtain unbiased estimates, and has introduced a method for estimating of the extent of potential over-treatment.

Our findings may contribute to a better scientific understanding of viremia and its rapid clearance in the first 3 months. It remains unclear whether these are transient infections or whether they represent the presence of non-replicating viral fragments. Our results and methods may also be relevant to other vertically acquired infections such as hepatitis B, where there are similar complexities introduced by clearance and the uncertain significance of a single positive virological marker in the exposed newborn.[34, 35]

More accurate information on both vertical transmission, clearance rates, and time to detectable RNA will become available when large scale randomized trials of early treatment or treatment in pregnancy are undertaken. In the meantime, routine testing of anti-HCV in antenatal or neonatal samples, now recommended in the USA[36] and shown to be cost-effective at a prevalence of 0.07%,[37] would allow HCV-RNA positive infants to be identified and followed until treatments become available.

## Data Availability

The data analysed for this paper was secondary. Individual patient data were requested from the studies listed in the manuscript and must be requested from the original research teams.

## Notes

### Author contributions

AEA conceived and carried out the analyses with the assistance of FG. AEA wrote the first and subsequent drafts of the paper. KS carried out the literature search and review. AEA, AJ, JC, and DMG were co-investigators on the HCVAVERT project, and AJ was the principal investigator. EC was a researcher on the HCVAVERT project. LP, EM-B, DMG and KB were senior or principal investigators on the 3 contributing studies: EPHN (European Pediatric HCV Network); ALHICE (Alpes-Maritimes, Languedoc, Haute Garonne Infection C chez l’Enfant); BPSU (British Pediatric Surveillance Unit). GI provided clinical input on hepatology and management of pediatric HCV. Curation of the original data files available to the project was the responsibility of LP and CT (EPHN), DMG, JC and KB (BPSU), and EM-B (ALHICE). Subsequent data processing was by FG and AEA. All authors critically reviewed and revised drafts as necessary, and approved the final version for submission.

### Disclaimer

The funding sources did not have any influence on study design, data collection, analysis and interpretation of the data, writing of the report, or the decision to submit for publication.

## Acknowledgements

The authors express their gratitude to all those who have contributed to the EPHN, BPSU and ALHICE studies.

