## Supplementary Materials for "Spontaneous Clearance of Vertically Acquired Hepatitis C Infection: Implications for Testing and Treatment"

*Joint last authors

### A. LITERATURE REVIEW

#### *Literature search*

We searched Pubmed on 19th March 2021 using the search terms: ((hepatitis C[Title/Abstract]) OR (hcv[Title/Abstract])) AND ((natural history[Title/Abstract]) OR (clear*[Title/Abstract]) OR (vertical transmission*[Title/Abstract])) AND ((child*[Title/Abstract]) OR (infant[Title/Abstract]) OR (paediat*[Title/Abstract]) OR (pediat*[Title/Abstract]) OR (mother*[Title/Abstract]) OR (vertical transmission*[Title/Abstract]) OR (pregnan*[Title/Abstract])) NOT (child-pugh[Title/Abstract).

Study reports were included if they clearly described clearance of vertically-acquired HCV in untreated children, with a denominator, number clearing, and method of recruitment. Studies were excluded if the vertical transmission rate was based on less than 20 mothers-child pairs, and or if clearance was based on less than 5 HCV infected infants. Only studies published from 1998 onwards were included, as earlier studies tended to be less clearly described.

The review identified 21 studies (Fig A1), which fell into 3 categories (Tables A1, A2, A3). Nine studies reported acquisition and clearance of viremia defined as at least one positive RNA test (Table A1): transmission rates were in the range 12%-70%, and clearance rates 66%-100%. Five of the nine studies also reported results for confirmed infection, with VT rates 3.5%-13% and clearance rates 14%-75%.

**Figure A1. Systematic Review flow chart**

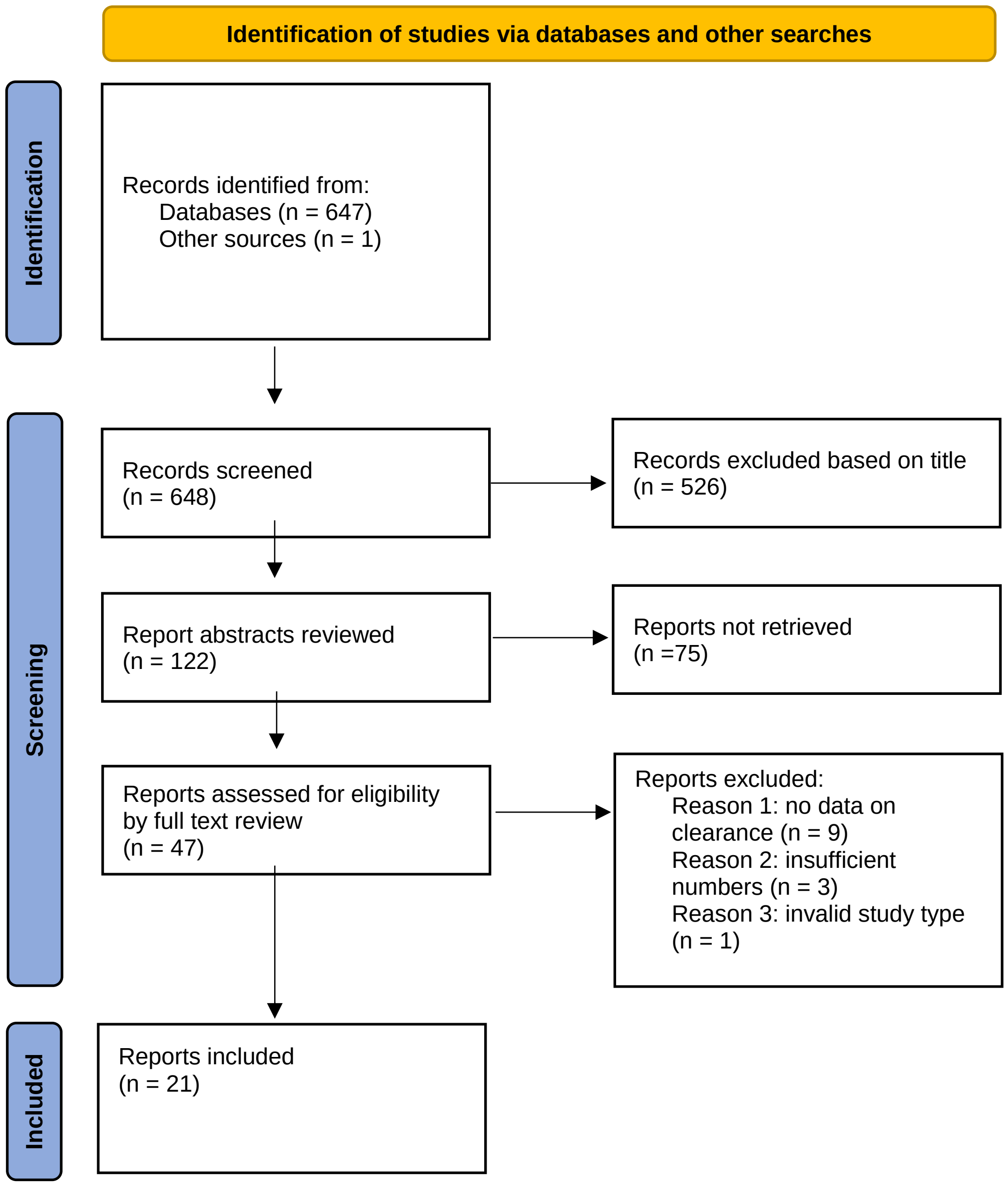

There was a second set of 5 studies in which infants were followed from birth but not always tested at delivery (Table A2). Some infections would therefore have cleared without ever being observed. These studies reported clearance rates of 13%-27%, based on analyses that took no account of delayed entry.  A third set of seven clinic-based studies followed children from referral: these reported 20%-30% clearance after 3 to 9 years follow-up, but did not account for delayed entry to the risk set (Table A3). These studies would have inevitably underestimated overall clearance as the majority of clearance would have already occurred prior to recruitment into the study. They are also likely to underestimate rates of clearance subsequent to referral and recruitment into the study, for two reasons: firstly, some studies only followed up patients who had not cleared for the initial 6-12 months they were under observation. Secondly, retrospective recruitment of babies for the purpose of further follow-up, even if they have been tested from birth, is likely to result in selective inclusion of children who are still under clinic care at the date when recruitment occurs, and exclusion of those who have already cleared and been discharged.

**Table A1.** Infants tested at delivery, VT rates and clearance in Confirmed and Suspected infection combined, and Confirmed infection only.

| | | Viraemia | | | Confirmed infection | | | |
|---|---|---|---|---|---|---|---|---|
| Lead author (year published) | Number mother-child pairs | VT rate | Number infected infants | **% cleared** | VT rate | Number infected infants | **% cleared** | By Age, years |
| Giacchino (1998)[1] | 45 | 20.0 | 9 | **67** | | | | 1.0 |
| Granovsky (1998)[2] | 122 | | | | 6 | 7 | **14** | 1.5 |
| Ruiz-Extremura (2000)[3] | 67 | 11.9 | 8 | **88** | | | | 1.5 |
| Ketzinal-Gilad (2000)[4] | 23 | 21.7 | 5 | **100** | | | | 0.25 |
| Ceci (2001)[5] | 60 | 50.0 | 30 | **93** | 13.3 | 8 | **75** | 1.0 |
| Mast (2005)[6] | 190 | | | | 4.7 | 9 | **33.3** | 2 |
| Shebl (2009)[7] | 232 | 12.5 | 29 | **72** | 6.5 | 15 | **47** | 5 |
| Ruiz-Extremura (2011)*[8] | 128 | 20.3 | 26 | **65.8** | | | | 6 |
| Bal (2016)[9] | 142 | 66.9 | 95 | **94.7** | 3.5 | 5 | **NR** | 1.5 |

* Positive RNA tests were confirmed within a few days.

**Table A2.** Infants followed from birth, but not necessarily tested at birth: delayed entry to risk set not taken into account. These studies reported confirmed infection only.

| Lead author (year published) | Number infected infants | **% cleared** | By Age, years | Comments |
|---|---|---|---|---|
| Tovo (2000)[10] | 104 | **13.5** | 4.1 | Inclusion conditional on regular follow-up |
| Resti (2003)[11] | 62 | **19** | 4.8 | Infants recruited between 3 and 12 months, follow-up conditional on infection. |
| Rerksuppaphol (2004)[12] | 16 | **13** | 8.7 | Mean age diagnosis 5 years |
| EPHN 3-broad (2005)[13] | 155 | **24** | 3 | ~40% retrospectively recruited infants enter study aged 2 – 6 months, inclusion conditional on regular follow-up[14] |
| Garazzino (2014)[15] | 45 | **27** | 12 | Inclusion only if anti-HCV at 18 months |

.

**Table A3.** Clinic-based studies: follow-up from referral.

| Lead author (year published) | Number of infected infants | Mean age at recruitment, years | Mean length of follow-up, years | Clearance, % | Comments |
|---|---|---|---|---|---|
| Jara (2003)[16] | 35 | NR | 6.2 | **17.1** | Only included if followed for > 1y |
| Iorio (2005)[17] | 66 | NR | 8.8 | **15** | 39% not vertically infected. Only included if anti-HCV positive for > 6 months |
| EPHN (2005)[13] | 85 | >0.5 | ~3 | **2.3** | Inclusion conditional on regular follow-up[14] |
| Bortolotti (2005)[18] | 270 | 0.25 | NR | **5.6** | 35% not vertically infected. Children with “early occurrence of viraemia clearance” excluded |
| Yeung (2007)[19] | 34 | >2 | 2.8 | **29** | Only included if followed for > 6 months |
| Bortolotti (2008)[20] | 227 | 3.4 | 5 | **11.4** | Only included if followed for > 1y |
| Mizuochi (2018)[21] | 348 | 3.1 | 7.8 | **8.6** | 10% not vertically infected. Only included if followed for > 1y |

### B. DATA SOURCES

**Table B1.** Period of recruitment, pediatric follow-up schedule, and risk factor distribution in the three cohorts. EPHN European Pediatric HCV Network, BPSU British Pediatric Surveillance Unit, ALHICE Alpes-Maritimes, Languedoc, Haute Garonne Infection C chez l'Enfant. Infants with no laboratory follow-up data were excluded from the table.

| | | **EPHN** | | **BPSU** | | **ALHICE** | |
|---|---|---|---|---|---|---|---|
| **Period of recruitment** | | 1999-2004 | | 1994-1999 | | 1999-2003 | |
| **Follow-up** | | Birth, 6w, 3m, 6m, 9m, 12m, 18m, 24, then every 6m | | Birth, 6w, 3m, 6m, 9m, 12m, 18m, 24, then every 6m | | Birth, 3m, 6m, 12m 18m, 24m | |
| | | **N** | **%** | **N** | **%** | **N** | **%** |
| **Total sample size** | | 1551 | 100 | 444 | 100 | 214 | 100 |
| **Mother HIV** | + | 212 | 13.7 | 22 | 5.0 | 55 | 25.7 |
| | **-** | 1233 | 79.5 | 328 | 73.8 | 157 | 73.3 |
| | **NK** | 106 | 6.8 | 94 | 21.1 | 2 | 1.0 |
| **Mother PCR** | + | 469 | 30.2 | 0 | 0 | 138 | 64.5 |
| | **-** | 184 | 11.9 | 0 | 0 | 62 | 29.0 |
| | **NK** | 898 | 57.9 | 444 | 100 | 14 | 6.5 |
| **Mode of Delivery** | **ECS** | 453 | 29.2 | 355 | 6.1 | 45 | 21.0 |
| | **Not-ECS** | 1063 | 68.5 | 27 | 80.0 | 169 | 79.0 |
| | **NK** | 35 | 2.3 | 62 | 14.0 | 0 | 0 |
| **Breastfeeding** | **Yes** | 434 | 28 | 31 | 3.8 | 3 | 1.4 |
| | **No** | 999 | 64.4 | 396 | 89.2 | 173 | 80.8 |
| | **NK** | 118 | 7.6 | 17 | 7.0 | 38 | 17.8 |

**NK: Not known. ECS: Elective caesarean section**

### C. DEFINITIONS

#### *Infection status*

*Confirmed infection:* Infants were considered infected if they had at least two positive RNA tests (not necessarily consecutive) or anti-HCV positive after 18 months.

*Viraemia:* Children with a single positive RNA test were considered to have viraemia, unless they had negative RNA tests when aged > 6weeks, in which case the single positive RNA results were regarded as false positives.

#### *Clearance of infection*

*Clearance:* Children were only considered to have cleared if their last RNA test was negative, or in the absence of any RNA tests their last anti-HCV test was negative. Only a single marker of clearance was required, either a negative RNA test or a negative anti-HCV test. Clearance was considered to have occurred between the date of the first of the two negative markers and the date of the immediately preceding positive RNA test. For children who had only a single marker of clearance, the date of clearance was between their last negative RNA test and the preceding RNA positive test.

Of the 106 children with confirmed infection, 36 cleared and 27 of these (75%) had two markers of clearance. Of the 179 with viremia, 87 cleared, 81% of who had two markers of clearance

### D. STATISTICAL ANALYSIS OF CLEARANCE

#### *Likelihood for interval censored and left truncated data*

We extracted age at first RNA +ve test $a_i$, which is the age when individual $i$ enters the risk set; age at last RNA +ve prior to clearance, $l_i$; and age at first RNA -ve after clearance, $r_i$. If $S(t)$ is the survival curve (proportion remaining infected) at age $t$, then the likelihood for an infant $i$ who is observed to clear is $L_i = \left(S(r_i) - S(l_i)\right) / S(a_i)$. For observations right censored at $l_i$ the likelihood is $L_i = S(l_i) / S(a_i)$.

We fitted natural cubic splines.[22] Knots were placed at the midpoint of the last positive and first negative RNA tests, that is $(l_i + r_i)/2$.[23] Boundary knots were placed at the smallest and largest midpoints, and two interior knots were placed at the 33rd and 67th centiles of the midpoints. Because spline models can be unstable, Weibull, lognormal and loglogistic models with a "cure" parameter[24] were fitted as sensitivity analyses, to verify that the cubic spline results were sound. The cure parameter represents the proportion of infected children who (hypothetically) never clear.

***Overtreatment and incidence estimates***

We estimated four measures of "over-treatment", assuming a 100% effective treatment administered at age $t$

1. Proportion of all infections that would clear spontaneously after $t$ months if there was no further clearance after 60 months
2. Proportion of all infections that would clear spontaneously after $t$ months if there was 25% additional clearance after 60 months
3. Proportion of all treated infections that would clear spontaneously after $t$ months if there was no further clearance after 60 months
4. Proportion of all treated infections that would clear spontaneously after $t$ months if there was 25% additional clearance after 60 months

These were calculated respectively as:

$$F_1(t) = \left(S(t) - S(60)\right) / \left(1 - S(60)\right)$$
$$F_2(t) = \left(S(t) - S(60)*0.75\right) / \left(1 - S(60)*0.75\right)$$
$$F_3(t) = \left(S(t) - S(60)\right) / S(t)$$
$$F_4(t) = \left(S(t) - S(60)*0.75\right) / S(t)$$

An approximate mid-period incidence was calculated from weekly estimates of the proportion remaining infected, $S_w$:

$$\lambda_w = 2(S_{w-1} - S_w) / (S_{w-1} + S_w)$$

***Estimation***

Estimation was by Bayesian Markov chain Monte Carlo, using WinBUGS 1.4.3.[25] Cubic spline models converged within 40-60,000 iterations; posterior summaries were based on 400,000 iterations, 80,000 from each of 5 chains, after a burn-in of 80,000.With parametric cure models convergence occurred within 5,000-7,000 iterations, and posterior summaries were based on 120,000 iterations, 40,000 from each of 3 chains, after a "burn-in" of 20,000.

***Specificity-adjustment sensitivity analysis: sensitivity and specificity of RNA tests***

The likelihood was adjusted $L_i = p(I_i = 1)\left(S(r_i) - S(l_i)\right) / S(a_i) + p(I_i = 0)$ for clearers, and $L_i = p(I_i = 1)S(r_i) / S(a_i) + p(I_i = 0)$ for non-clearers, where $I_i$ indicates whether individual $i$ is infected, $I_i = 1$, or not $I_i = 0$. If $\theta_i$ is the probability that an individual is infected conditional on their test results – ie their positive predictive value, then $I_i \sim Bernoulli(\theta_i)$. Distributions for $\theta_i$, the Positive Predictive Value of RNA tests, were sourced as follows.

**Table D1.** Source of values for positive and negative predictive value of RNA tests

| **Parameter** | **Data** | **Distribution or calculation** | **Estimate (95%CrI)** | **Source** |
|---|---|---|---|---|
| Prevalence of vertically-acquired infection | [(106+179/2]/1749 | P1~Beta(142.5,1606.5) | 0.0815 (0.69-0.095) | Three cohorts (see text below) |
| Proportion of infections RNA+ve in first 3 days | 17/54 | W~Beta(17,37) | 0.31 (0.20-0.44) | Mok[26] |
| Prevalence in first 3 days | | P2 = P1 * W | 0.026 (0.016-0.037) | |
| Sensitivity | 165/177 | Se ~ Beta(165,12) | 0.93 (0.89-0.93) | Three cohorts (see text below) |
| False positive rate | 35/2495 | Fp ~ Beta(35,2460) | 0.014 (0.010-0.019) | Three cohorts (see text below) |
| PPV after 3 days | | PPV1 = Se*P1/(Se*P1+Fp*(1-P1) | 0.63 (0.50-0.75) | |
| PPV first 3 days | | PPV2 = Se*P2/(Se*P2+Fp*(1-P2) | 0.85 (0.81-0.90) | |
| NPV after 3 days | | NPV1=(1-Fp)*(1-P1)/[(1-Fp)*(1-P1)+(1-Se)*P1 | 0.994 (0.990-0.997 | |
| NPV after 3 days | | NPV2=(1-Fp)*(1-P2)/[(1-Fp)*(1-P2)+(1-Se)*P2] | 0.998 (0.997-0.999) | |

*Prevalence*. Average of the prevalence of confirmed infection and viremia.

*Sensitivity*. Denominator: number of RNA tests among those with confirmed infection which post-dated their first positive test and predated their last positive (177). Numerator: number of these that were positive (165) of these were positive.

*False positive rate*. Denominator: number of tests among those with confirmed infection that postdated their first negative after 6 weeks or which postdated 2 negative tests indicating clearance (2495). Numerator: number of those that were positive (35)

Our estimates of PPV are in line with previously published estimated,[27] except the latter were based on diagnostic sensitivity and specificity, whereas we have attempted to estimate analytic sensitivity and specificity. Estimates of Negative Predictive Value are also consistent with previous research.[27, 28]

### E. SENSITIVITY ANALYSIS: RESULTS

Two sensitivity analyses were carried out on both viraemia and confirmed infection: first we looked at sensitivity to model choice (cubic spline or parametric cure models); second, we looked at the applied the preferred model to a dataset in which clearance was based on two markers (Tables E1, E2). A third sensitivity analysis investigated a specificity-adjusted survival curve which allowed for the possibility of false positive RNA results in cases of viraemia based on just a single RNA positive test (Table E1).

#### *Viremia*

**Table E1.** Sensitivity analyses of clearance of viremia. Posterior mean deviance, median age (months) at clearance among those who clear within 6 months, and percent clearance. * Base-case model

| *Model* | *Posterior mean Deviance* | *Median age at clearance* | *Percent remaining not cleared* | | | | |
|---|---|---|---|---|---|---|---|
| | | | *3m* | *6m* | *18m* | *36m* | *60m* |
| ***Alternative survivals model, one marker of clearance*** | | | | | | | |
| **Cubic Spline** | **383.4** | **1.2 (0.1-2.7)** | **37.1 (19.2-54.7)** | **27.5 (14.2-41.0)** | **17.0 (8.5-26.1)** | **12.2 (5.8-19.8)** | **9.4 (4.1-16.5)** |
| Weibull | 381.7 | 0.6 (0-1.9) | 31.2 (14.4-49.2) | 23.4 (10.6-37.4) | 13.9 (6.4-22.7) | 10.6 (4.7-18.0) | 9.2 (3.8-16.6) |
| Lognormal | 380.9 | 0.7 (0-2.6) | 31.3 (12.9-49.9) | 23.6 (9.7-38.0) | 14.6 (6.1-24.0) | 11.0 (4.5-18.6) | 9.2 (3.6-16.3) |
| Loglogistic | 381.3 | 1.2 (0.2-2.7) | 38.5 (21.6-55.1) | 28.5 (15.9-41.7) | 17.5 (9.7-26.2) | 13.3 (7.2-20.6) | 11.2(5.8-18.2) |
| ***Two markers of clearance*** | | | | | | | |
| Cubic Spline | - | 1.5 (0.1-3.2) | 43.8 (23.7-62.0) | 33.0 (17.8-47.5) | 22.3 (11.8-33.1) | 18.2 (9.2-28.0) | 15.5 (7.4-25.1) |
| ***Specificity adjusted estimates*** | | | | | | | |
| Cubic Spline | - | 1.1 (0-3.1) | 45.5 (24.8-63.3) | 34.6 (19.0-49.4) | 21.9 (11.5-32.5) | 15.7 (7.8-24.7) | 11.9 (5.3-20.4) |

*Confirmed infection*

**Table E2.** Sensitivity analysis of clearance of confirmed infection. Posterior mean deviance, median age (months) at clearance among those who clear within 5 years; percent remaining infected at 3, 6, 18, 36 and 60 months; percent (95%CrI) of all infection that clears spontaneously after 36m assuming no further clearance after 60m. * Base-case model

| *Model* | *Posterior mean Deviance* | *Median age at Clearance* | *Percent Clearance* | | | *Percent of all infections that clear spontaneously after 36m, assuming no further clearance after 5 years* |
|---|---|---|---|---|---|---|
| | | | *12m* | *36m* | *60m* | |
| ***Alternative survivals model, one marker of clearance*** | | | | | | |
| **Cubic Spline *** | **228.6** | **12.4 (7.1-18.9)** | **67.4 (53.8-78.3)** | **42.7 (30.2-55.3)** | **34.1 (18.4-49.9)** | **12.8 (3.8-23.1)** |
| Weibull | 227.3 | 10.1 (4.0-16.4) | 63.6 (48.5-75.7) | 41.3 (29.3-53.7) | 34.7 (21.2-49.5) | 9.8 (2.0-19.1) |
| Lognormal | 227.8 | 10.5 (4.2-16.3) | 65.2 (50.5-76.6) | 43.8 (32.1-55.9) | 35.9 (23.8-49.3) | 12.3 (5.9-18.8) |
| Loglogistic | 227.4 | 12.0 (6.5-17.6) | 67.8 (55.3-78.4) | 44.0 (32.6-55.8) | 35.8 (23.6-49.4) | 12.6 (5.8-19.7) |
| ***Two markers of clearance*** | | | | | | |
| Cubic Spline | - | 11.6 (6.3-19.9) | 71.6 (58.7-1.9) | 53.7 (39.8-66.6) | 46.1 (28.1-62.6) | 13.7 (3.6-25.2) |
